# Capturing the implications of residential segregation for the dynamics of infectious disease transmission

**DOI:** 10.1101/2024.06.26.24309541

**Authors:** Jon Zelner, Danielle Stone, Marisa Eisenberg, Andrew Brouwer, Krzysztof Sakrejda

## Abstract

Occupational and residential segregation and other manifestations of social and economic inequity drive of racial and socioeconomic inequities in infection, severe disease, and death from a wide variety of infections including SARS-CoV-2, influenza, HIV, tuberculosis, and many others. Despite a deep and long-standing quantitative and qualitative literature on infectious disease inequity, mathematical models that give equally serious attention to the social and biological dynamics underlying infection inequity remain rare. In this paper, we develop a simple transmission model that accounts for the mechanistic relationship between residential segregation on inequity in infection outcomes. We conceptualize segregation as a high-level, fundamental social cause of infection inequity that impacts both who-contacts-whom (separation or preferential mixing) as well as the risk of infection upon exposure (vulnerability). We show that the basic reproduction number, ℛ_0_, and epidemic dynamics are sensitive to the interaction between these factors. Specifically, our analytical and simulation results and that separation alone is insufficient to explain segregation-associated differences in infection risks, and that increasing separation only results in the concentration of risk in segregated populations when it is accompanied by increasing vulnerability. Overall, this work shows why it is important to carefully consider the causal linkages and correlations between high-level social determinants - like segregation - and more-proximal transmission mechanisms when either crafting or evaluating public health policies. While the framework applied in this analysis is deliberately simple, it lays the groundwork for future, data-driven explorations of the mechanistic impact of residential segregation on infection inequities.

## 1 Introduction

Occupational and residential segregation, racial capitalism (1), mass incarceration (2) and other manifestations of social and economic inequity (3) have been definitively shown to drive of racial and socioeconomic inequities in infection, severe disease, and death from a wide variety of infections including SARS-CoV-2 (4,5), seasonal and pandemic influenza (6), tuberculosis (7,8), STIs (9) and many others.

Despite a deep and long-standing quantitative and qualitative literature on infectious disease inequity, mathematical models that give equally serious attention to the social and biological dynamics underlying infection inequity have been in short supply. This has had serious consequences for the ability of public health authorities to anticipate and respond to infection inequality: For example, a suite of mathematical modeling tools was ready to be pressed into action at the beginning of the COVID-19 pandemic. But the absence of models ready to address the inequities in SARS-CoV-2 infection, severe disease and death that emerged almost immediately highlighted a large and dangerous blindspot in pandemic preparedness, as well as in the day-to-day application of transmission models in infectious disease epidemiology (3).

While the uptick in equity-oriented modeling in the wake of the pandemic is a positive development (10), it is essential that these models treat social mechanisms with the same theoretical and empirical rigor that they bring to biological ones (11). Otherwise, we risk encoding old assumptions with deep roots in scientific racism, e.g. about increased susceptibility of Black people to infection and death from a range of infections due to genetic inferiority, poor adaptation to urban environments (7), and more recently, an assumed high prevalence of comorbid infections related to stigmatized health behaviors (12) rather than differential exposure associated with segregation and other forms of discrimination (14).

The increased influence and high visibility of infectious disease modeling as a field during COVID-19 also creates the risk of ‘engineer’s syndrome’ in which perceived success in solving one set of problems increases our confidence - and hubris - with respect to solving other ones for which our existing tools are ill-suited. The risk of harm from this - even if well-intended - is very real: In addition to providing an inaccurate characterization of population-wide and group-specific disease dynamics, models informed primarily by bias-inflected ‘common sense’ about the causes of infection inequity risk further stigmatizing the communities they purport to be characterizing and, by implication, are trying to help (16).

### 1.1 Modeling the fundamental causes of infection inequity

A principled applciation of fundamental cause theory (FCT) (17) to the problem of infectious disease transmission has been suggested by multiple authors as a useful theoretical anchor for mathematical models of infectious disease inequity (3). In this framework, phenomena such as socioeconomic inequality, social stigma (19), structural racism (20), and residential segregation (21) function as high-level ‘fundamental’ causes of infection inequity that orchestrate the down-stream, more-proximal processes of exposure, infection, illness, and death (17). The present analysis represents an effort to employ the ‘systems of exposure’ approach to FCT advocated for by Riley (22) and others in the context of infectious disease transmission modeling.

Residential segregation provides a good starting point for developing better equity-oriented transmission models for a number of reasons: 1) Segregation is a well-recognized driver of health inequity, 2) There is a deep social science literature on the topic, and 3) Segregation is understood to have impacted both risks of transmission and access to care, testing, and other relevant services during COVID-19 pandemic. In addition, from an intervention perspective, residential segregation is an important and modifiable dimension of race/ethnic and economic inequity (23) in infection risk that can be addressed by activism and public policy.

Despite a dearth of fully-fledged models linking residential segregation to infection inequity, mechanistic linkages between segregation and infection inequity have been explored in great depth by many other authors. For example, in his paintstaking dissection of racial inequity in Tuberculosis infection and mortality in Jim Crow era Baltimore, the medical historian Samuel Kelton Roberts (7) illustrated the multiplex connections between institutionalized segregation and racism and TB infection and death. This included not only differential rates of exposure and treatment by the medical establishment, but also the wholesale manipulation of the housing code to exclude the types of dwellings Black residents were consigned to from regulations to limit crowding, and improve ventilation and fire safety.

In an influential paper, Acevedo-Garcia (24) framed these relationships in terms of the measures of residential segregation typically used in social demography and hypothesized relationships between various measures of residential segregation and the mechanisms by which segregation impacts infectious disease transmission. In the conceptual model originally outlined in (24) the world is broken into two groups: an advantaged majority and a disadvantaged minority. This generic framing is important. Although residential segregation in the United States is often imagined in racial or socioeconomic terms, in the context of infectious disease transmission, we posit that the literature on residential segregation should inform our thinking about patterns of contact and infection risk in any setting where there are two or more interacting populations with differing rates of *within* and *between-group* contact.

Owing to the long history of institutionalized segregation in the United States from the Jim Crow era to 20th-century redlining and racial covenants, and their ongoing present-day impacts (25), in the U.S. residential segregation is often reflexively conceptualized primary in terms of race and ethnicity. However, segregation can - and does - occur globally along many other dimensions including religion (26) or at the intersection of class and race (27). In contexts of large-scale, institutionalized segregation, such as the Jim Crow-era United States, apartheid-era South Africa (28) or present-day occupied Palestine, patterns of high-intensity intra-group exposure are driven by concrete policies and physical infrastructure. But these mechanisms may also be more informal, reflecting the persistent physical and social residues of institutionalized discrimination.

### 1.2 Residential segregation as a bundle of risks

In the essay *Race as a Bundle of Sticks*, Sen & Wasow (23) outline a theoretical and methodological approach to capturing the causes of racial differences in political behavior from a constructivist perspective, i.e. one in which the ‘effects’ of race as not located in the racial category itself (i.e. the ‘essentialist’ perspective) but instead in the constellation of socio-structural processes that are attached to race. Our goal is to apply this line of thinking to residential segregation.

The diagram in Figure 1 (adapted from (24)), illustrates how high-level causes like structural racism and discrimination produce residential segregation. Segregation is then hypothesized to drive infection inequity via its impact on multiple more proximal mechanisms impacting the on-the-ground transmission proces. In this analysis we examine the individual and joint implications of two of the most critical dimensions of residential segregation on the social patterning of acute infection risk. Specifically, we focus on:

**Figure 1.**
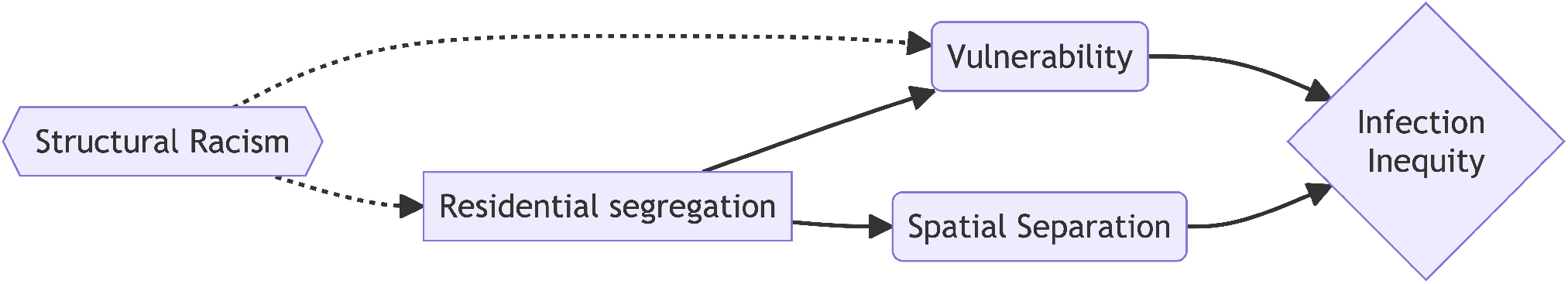
Flow diagram relating the indirect impact of residential segregation on infection inequity via specific dimensions of segregation. Solid and dashed lines represent hypothesized mechanisms included and not included in the present model, respectively.

1. **Spatial separation:** This is often used as a proxy measure of who-contacts-whom. For example, individuals living in the same neighborhood are often assumed to contact each other at a higher rate than those living in different neighborhoods. During the COVID-19 pandemic, mobility data were used to illustrate in more detail the relationship between residential location and exposure risk in the community. For the purposes of this theoretical analysis, we will conflate spatial location and group-specific contact.
2. **Vulnerability:** Our operationalization of vulnerability is meant to capture an increased risk of infection upon exposure for individuals belonging to a marginalized or disadvantaged group that is subject to segregation. This stylized representation is meant to reflect increased *intensity* of exposure, e.g. owing to living in crowded or poorly ventilated housing, working in high-exposure settings (e.g. home healthcare, food service), and potentially increased susceptibility to infection due to exposure to environmental contaminants, household and workplace environments characrerized by crowding and poor ventilation, or due to comorbid health conditions that may increase vulnerability.

The specific goal of the modeling exercise in this paper is to understand how these high-level social mechanisms, working separately and in concert with each other, drive inequity in infection outcomes. Capturing the interaction between vulnerability and separation is essential. This is because a key tenet of FCT is that the intermediary mechansisms driving inequality in disease outcomes will necessarily be correlated with each other because they share a common upstream cause, i.e. structural racism. In other words, we should generally expect increased vulnerability to accompany a higher degree of spatial separation, even when one doesn’t directly cause the other. For this reason, the point of studying their independent effects on group-specific and population-wide risks is to understand the relative contribution of each sub-mechanism to the total effect of residential segregation on infection inequity.

We develop a more comprehensive understanding of the relationships between these basic mechanisms and infection inequity by interrogating a mathematical model connecting the mechanisms illustrated in Figure 1. It is important to recognize, however, that the model discussed in this analysis represents a small subset of the processes that generate differential infection outcomes, not to mention rates of severe disease and death(13).

## 2. Methods

In this section, we will first describe the mechanistic transmisison model at the heart of our analysis, and then the analytic and simulation-based tools we will use to analyze the behavior of this model.

### 2.1 Transmission Model

We adapted the model used by Ma et al. (5) to analyze race-ethnic differences in early COVID-19 mortality in the New York City area. This model is based on the preferential mixing model originally described by Hethcote (29). In our version of this model, we define a parameter *ϵ* which varies from 0 to 1 and represents the proportion of time an individual spends mixing randomly with all members of the population (including members of their own group) as opposed to with only members of their own group. In our implementation of this model, *ϵ* = 0 is equivalent to a homogeneously mixed population and *ϵ* = 1 is analogous to a fully segregagted one, i.e. where thereis no crossover in contact beteween the population groups.

#### Spatial separation

The preferential mixing parameter, *ϵ* can be understood as being analogous to the index of dissimilarity, which is often used to measure the intensity of residential segregation. Dissimilarity is typically understood as a measure of evenness in the distribution of individuals (or their attributes) across geographic areas. A low value of dissimilarity, e.g. 0, indicates that no one needs to move to a new location in order to facilitate mixing proportional to the size of each population group. By contrast, a dissimilarity value close to 1 indicates that nearly all of the individuals in one of the groups would have to move to another location in order to acheive an even, proportional distribution of individual attributes across locations.

#### Vulnerability

The relative vulnerability of the minority group as compared to the majority group is represented by the parameter *ρ*, which scales the force of infection experienced by members of the disadvantaged group upwards. This parameter is allowed to take on values ≥ 1, with larger values representing increased vulnerability to infection.

We explored two specifications of vulnerability: One in which individuals are only more vulnerable to exposures to other members of the minority group, and another in which minority group members are more vulnerable to all exposures regardless of their source (i.e. minority or majority group). Because there were only very small quantitative differences in our measured outcomes for these different model specifications, we focus here on the latter model - increased vulnerability to all contacts - for the sake of clarity. Equation 1 shows how the mechanisms of spatial separation and vulnerability are combined to calculate the force of infection.

Equation 2 below defines a susceptible-infected-susceptible (SIS) differential equation system in which the rates of infection for each group are a function of the preferential mixing parameter (*ϵ*) and relative vulnerability to infection (*ρ*):

**Table 1:**
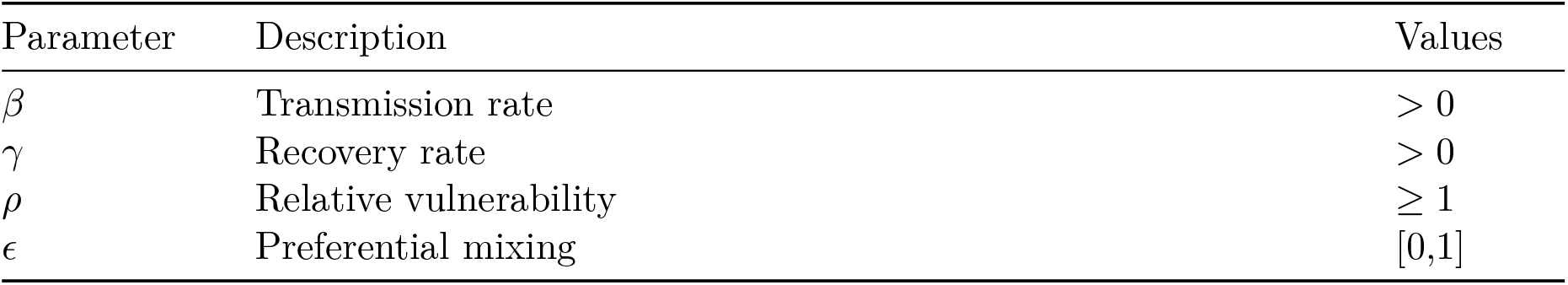
Key model parameters.

| Parameter | Description | Values |
| --- | --- | --- |
| $\beta$ | Transmission rate | $> 0$ |
| $\gamma$ | Recovery rate | $> 0$ |
| $\rho$ | Relative vulnerability | $\geq 1$ |
| $\epsilon$ | Preferential mixing | $[0,1]$ |

First, we will write the force of infection for each group (λ_1_ & λ_2_) as follows:

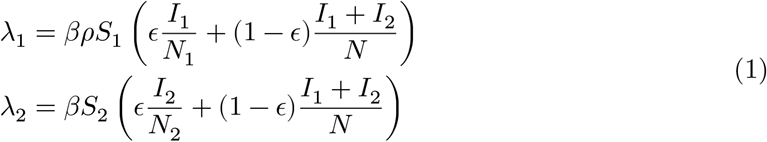

We can then include these in an SIS model framework, assuming equal durations of infectiousness for each group:

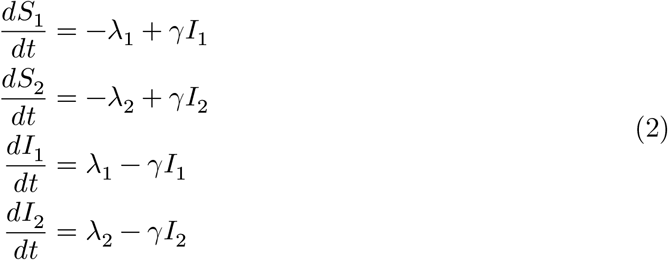

We utilize an SIS-type system to capture trasnmission dynamics of a stylized *endemic* infection in which reinfection is possible. This is not meant to represent a specific pathogen, which would have a more complex natural history, but instead to provide a way of thinking about how the mechanisms associated with segregation impact the behavior of this commonly-employed system.

## 3. Results

In this section, we first derive an expression for the basic reproduction number (ℛ_0_) as a function of the key parameters of the model described in Equation 2. We then utilize numerical and analytic results from the model to understand how different parameter combinations result in differing distributions of infection between the minority (*I*_1_) and majority (*I*_2_) groups. All symbolic analysis was conducted using s(30) in Python 3.11. Numerical simulation from differential equation models was completed using scipy (31). Figures were generated using plotnine 0.12.4 (32) for Python.

### 3.1 Deriving ℛ_0_ as a function of prferential mixing and differential vulnerability

In this section, we walk through the steps outlined by Brouwer (33) to derive an expression for ℛ_0_ for this model using the next-generation method presented in (34) and others. We will walk briefly through the steps outlined in (33), but interested readers should refer to this manuscript for a detailed explanation.

#### Next generation method

We first note that 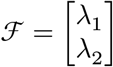 is the vector of rates at which previously uninfected people enter the infectious compartments. Per the next generation method, we calculate the Jacobian of ℱ as a function of *I*_1_ and *I*_2_ near the disease-free equilibrium.

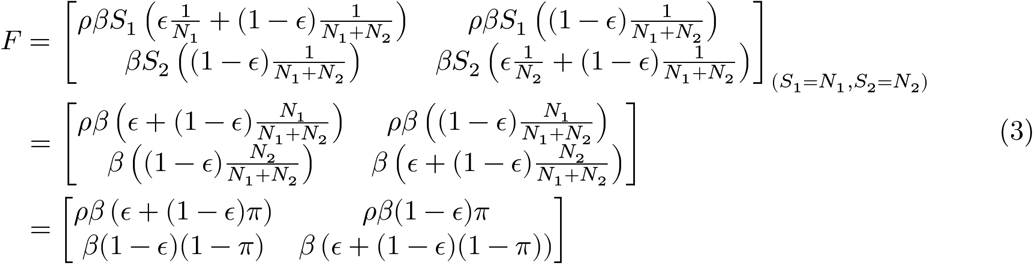

where we define *π* := *N*_1_/(*N*_1_+*N*_2_) as the fraction of the population in Group 1. The (*i, j*) entry of the *F* matrix is the rate at which an individual in group *j* creates infections in group *i*. The matrix *V* , which here denotes the Jacobian of the recovery rates at the disease-free equilibrium, is given simply by 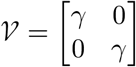 The next generation matrix is defined as *K* = *F V* ^−1^, and the entries (*i, k*) of *K* denote the expected number of new infections in group *i* given an infected individual in group *k*. Here,

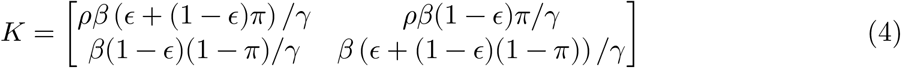

The basic reproduction number is defined as the spectral radius of *K*. For a two-group model, we have:

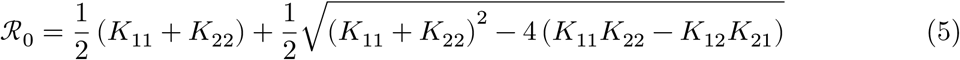

For our model, we can write:

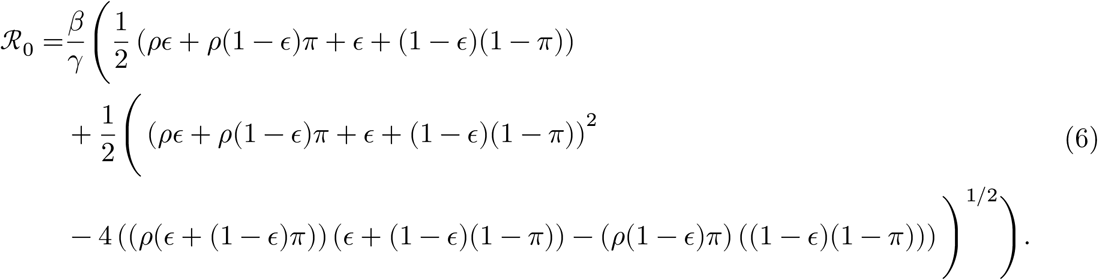

Although this expression appears quite complicated, we can build some intuition by looking at some simplifying scenarios. First, suppose we have *ρ* = 1 and *ϵ* = 0. In this instance, there is no vulnerable group, and the groups are well mixed. Then, it is not surprising that ℛ_0_ simplifies to *β*/*γ*. If *ρ* = 1 but 0 < *ϵ* ≤ 1, we have a situation with assortative mixing but no vulnerability, and ℛ_0_ again simplifies to *β*/*γ*. On the other hand, if we have *ρ* > 1 but *ϵ* = 0, so that one group is more vulnerable, but the groups are well mixed, ℛ_0_ simplifies to *β*(*ρπ* + (1 − *π*))/*γ* meaning that the basic reproduction number is an average of the basic reproduction numbers in the two groups, weighted by population size. Thus, it is the particular interaction of the vulnerability, social separation, and relative population size that leads to the formulation of ℛ_0_.

It may be helpful to understand this interaction by looking at concrete example. Let us assume a situation in which the groups mix roughly in proportion to their population share (*ϵ* = 0) and that group 1 has no extra vulnerability (*ρ* = 1). Let’s assume that the baseline transmission rate is *β* = 1.5, the recovery rate is *γ* = 1, and group 1 is *π* = 1/4 of the population. The next generation matrix now looks like this:

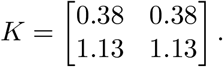

These values mean that one infection, regardless of group (as the columns are identical), will result in an expected 0.38 infections in Group 1 and 1.13 infections in Group 2. Here, ℛ_0_ = 1.5 and the eigenvector of K correpsonding to ℛ_0_, (0.25, 0.75), indicates that infections are generated proportionally to the population sizes of the groups.

Now, let’s assume that Group 1 is twice as vulnerable as Group 2 (*ρ* = 2). Then,

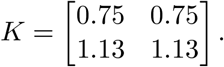

Now, one infection is expected to generated 0.75 infected people of group 1 instead of 0.38. Now, ℛ_0_ = 1.88 and the eigenvector of K correpsonding to ℛ_0_, (0.40, 0.60), indicates that more infections are generated in group 1 than is proportional.

Now, let’s further assume that mixing is highly assortative (*ϵ* = 0.9). Then, the balance of new infections swings dramatically so that most new infections are within group 1.

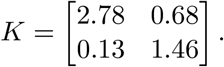

This change further increases ℛ_0_ = to 2.83, with an eigenvector of (0.92, 0.08) indicating that over 90% of new infections occur among group 1.

Figure 2 shows these these relationships in more detail, over a wider range of parameter values. Specifically, it illustrates how the value of ℛ_0_ changes for different values of *ρ* and *ϵ* and how these differences impact the relative distributions of infection at equilibrium for the minority and majority groups respectively.

**Figure 2.**
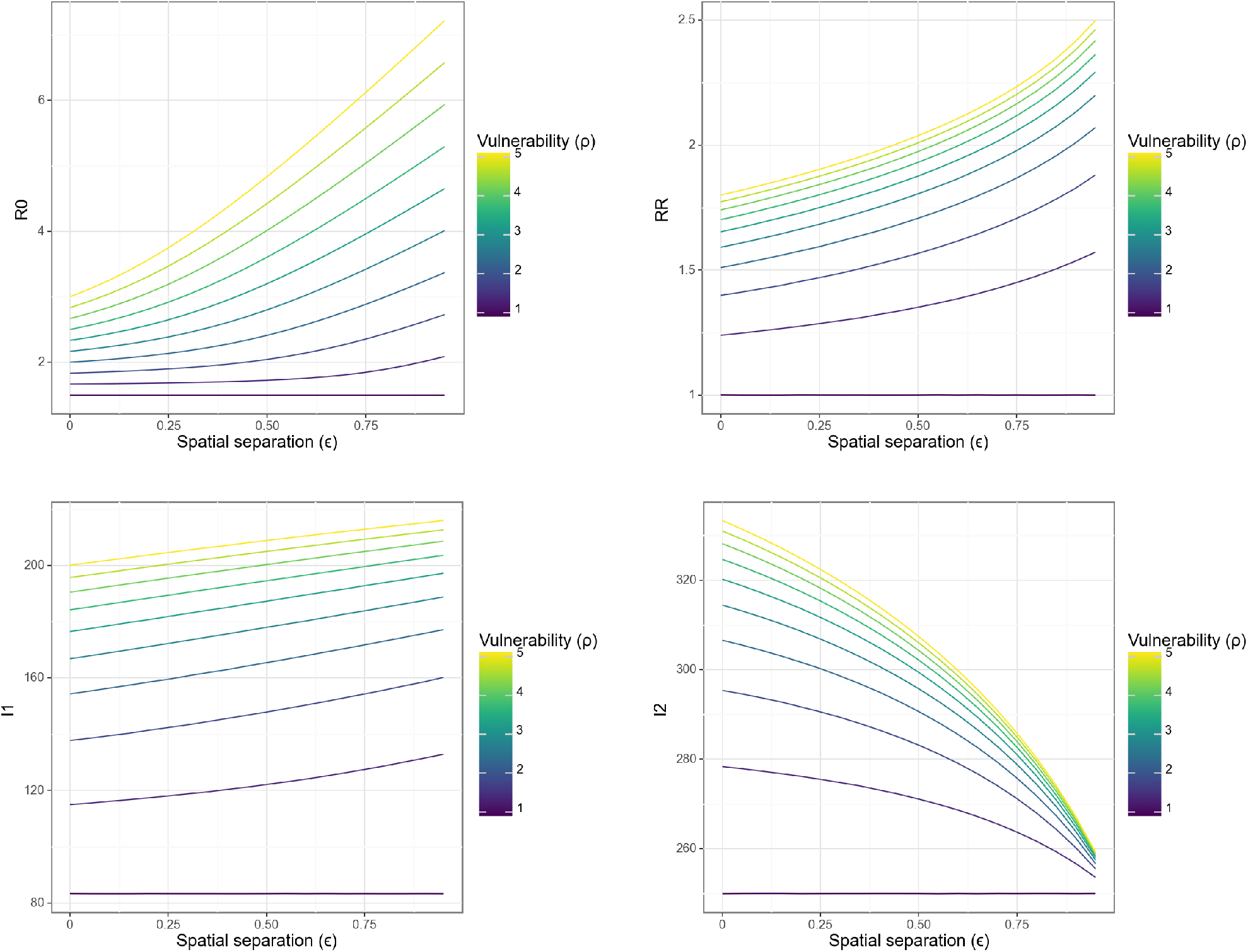
ℛ_0_ and relative risk of infection at equilibrium as a function of preferential mixing intensity (*ϵ*) and vulnerability to infection (*ρ*). The top left-hand panel illustrates the scaling of the population-level parameter ℛ_0_ as a function of increasing segregation. The top right-hand panel illustrates scaling of the relative risk of infection for the minority group vs. majority group as a function of increasing segregation. The bottom two panels represent the absolute prevalence in each group as a function of the model parameters. Each colored line represents a different value of the vulnerability parameter *ρ*. The fixed parameters are *β* = 1.5, *γ* = 1, *N*_1_ = 250 and *N*_2_ = 750.

### 3.2 Characterizing the stability of inequity

One way to think about the role of residential segregation in an infectious disease system is that it creates an equilibrium of inequity that a disease system returns to after some kind of perturbation. This may occur in a situation such as an emerging infection in which initial exposure is broad-based, but over time a transient period of shared risk converges towards a state of more stable, long-run inequity. Figure 3 illustrates what this may look for different combinations of input parameters.

**Figure 3.**
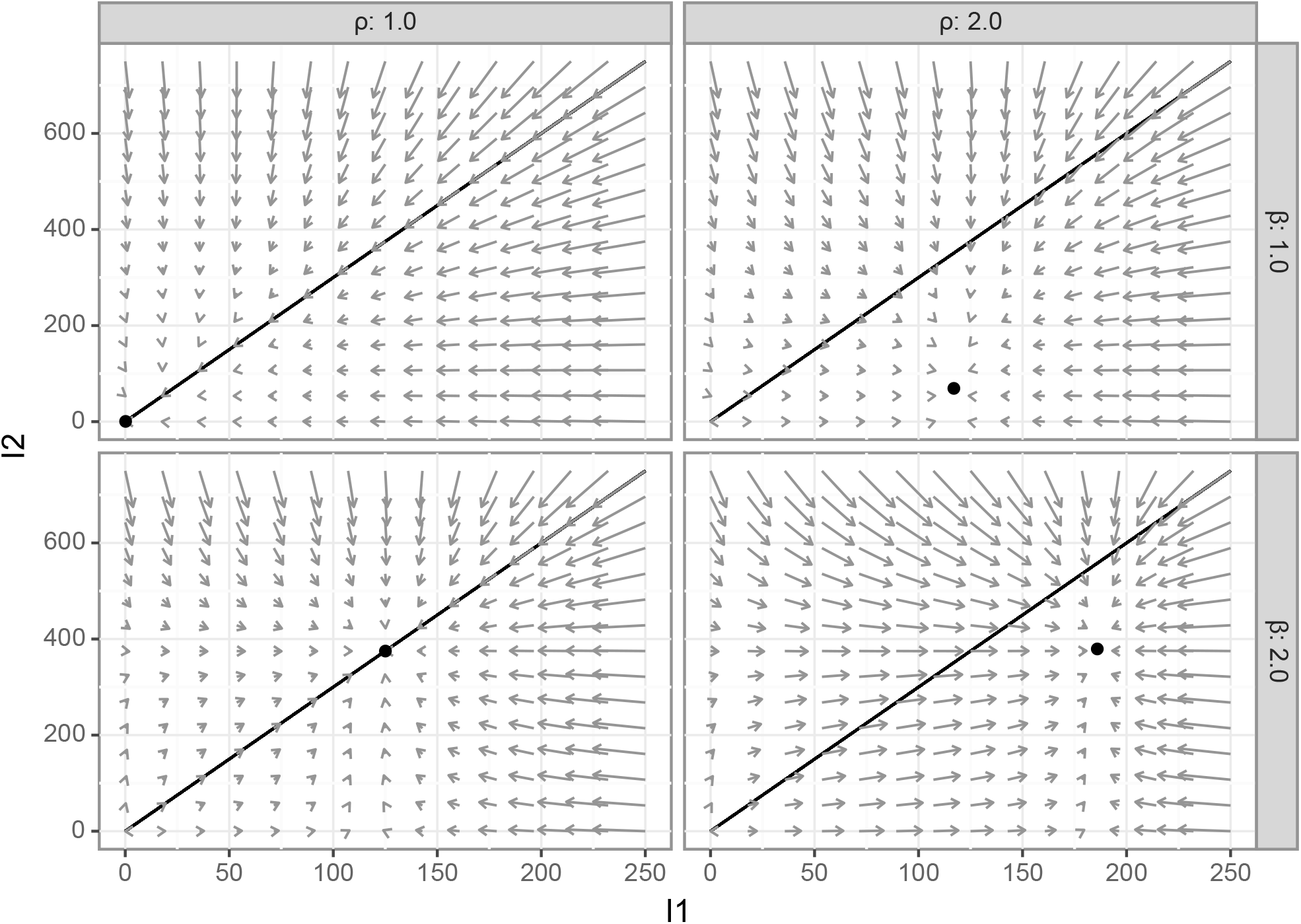
Phase diagrams illustrating several risk-inequity equilibrium scenarios under highly segregated contact (*ϵ* = 0.91). Each panel in the figure illustrates the phase diagram for a different combination of the baseline transmission rate (*β*) and the vulnerability parameter (*ρ*). The point in each panel shows the long-run equilibrium values of the prevalence of infection among the minority group (I1) and majority group (I2). Gray arrows indicate the direction and rate of change at each set of initial conditions. The diagonal line represents a scenario of equality in infection: values below the diagonal represent greater risk in the minority group, while values below indicate greater risk for the majority group.

For the panels in the left-hand column, risk is shared equally, with either only the diseasefree equilibrium available, equal per-capita prevalence (represented by the solid diagonal line). The right-hand panels show a scenario of increased vulnerability (*ρ* > 1) for the minority group. These show two important scenarios: In the topmost column, the outbreak goes from subcritical to critical for the entire population, but the long-run risk is greater for the minority than the majority group. In the bottom panel, an increase in vulnerability has a small effect on prevalence for the majority but a large one for the minority, illustrating the way that changes in vulnerability increase inequity for a given intensity of isolation and baseline transmission risk.

### 3.3 Modeling residential segregation as a fundamental cause of infection inequities

Until this point, we have examined the impacts of the two dimensions of residential segregation outlined by Acevedo-Garcia (24) on infection inequity as independent parameters. However, if we conceptualize the abstract process of segregation as a high-level fundamental cause which drives these intermediary mechaisms, we should expect them to be correlated with each other. In other words, fundamental cause theory would predict that, because they are both a function of high-level structural discrimination, spatial separation and vulnerability to infection should both increase as the intensity of the socio-legal regime of residential segregation increases.

In this section, we complete our analysis of segregation as a fundamental cause of infection inequity by introducing a correlation between preferential mixing, *ϵ* and vulnerability, *ρ*, as outlined in Figure 1. To do this, we introduce the parameter *τ* which represents the value of *ρ* when *ϵ* = 1, i.e. vulnerability under maximal preferential mixing.

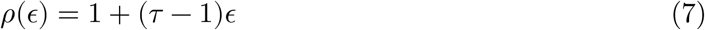

In the formulation in Equation 7, *ρ* = 1 when *ϵ* = 0, up to 1 + *τ* when *ϵ* = 1, i.e. there is no difference in vulnerability in the no-segregation scenario and it reaches its maximum under full separation. It is important not to interpret the relationship in Equation 7 as a causal one, i.e. in which physical separation induces greater vulnerability. Instead, we can think of the effect of *τ* as inducing a correlation reflecting a scenario in which greater physical separation is associated with greater vulnerability to infection among the minority population, reflecting the reality that separate is inherently unequal. Equation 8 shows the next-generation matrix for this model. Notably, the infection risk for the minority group is now proportional to *ϵ*^2^ when *τ* > 1.

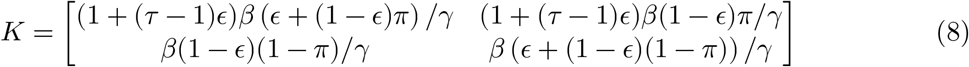

Equation 9 shows ℛ_0_ resulting from applying the two-group approach in Equation 5 to the elements of Equation 8. In comparison to the derivation of ℛ_0_ in Equation 6 where *ϵ* and *ρ* were allowed to vary independently, in the fundamental cause version of our model, ℛ_0_ = *β*/*γ* only when *τ* = 1 or when *τ* > 1 and *ϵ* = 0, i.e. there is no segregation. While the expression in Equation 9 is complex, the most important intuitive takeaway from it that ℛ_0_ is now a non-linear, increasing function of the parameter governing preferential mixing, *ϵ*.

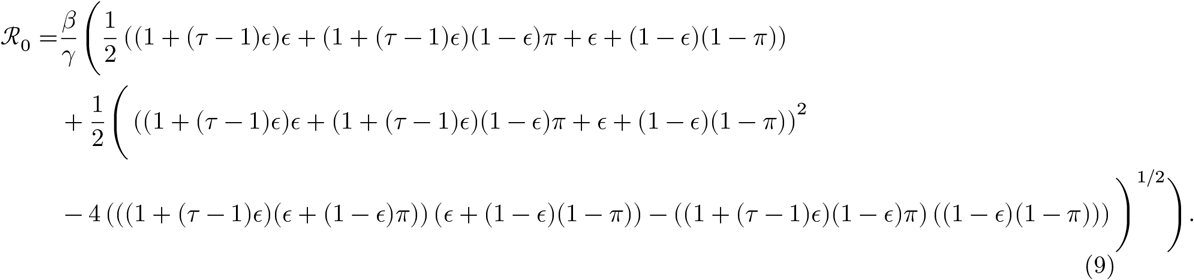

This is illustrated in Figure 4 which shows that in this setup, ℛ_0_ grows super-linearly with *ϵ*, and that the pace of this rise is faster for higher levels of *τ* . This figure also shows that the relative risk of infection for the minority group grows rapidly with *ϵ* for all *τ* > 1. Notably, however, with increasing segregation intensity, the burden of infection in the majority, advantaged group, eventually slows or declines, even as population-wide ℛ_0_ grows dramatically. This finding echoes those from (5) and illustrates the critical importance of understanding these dynamics from both a population-wide (ℛ_0_) and between-group distributional perspective.

**Figure 4.**
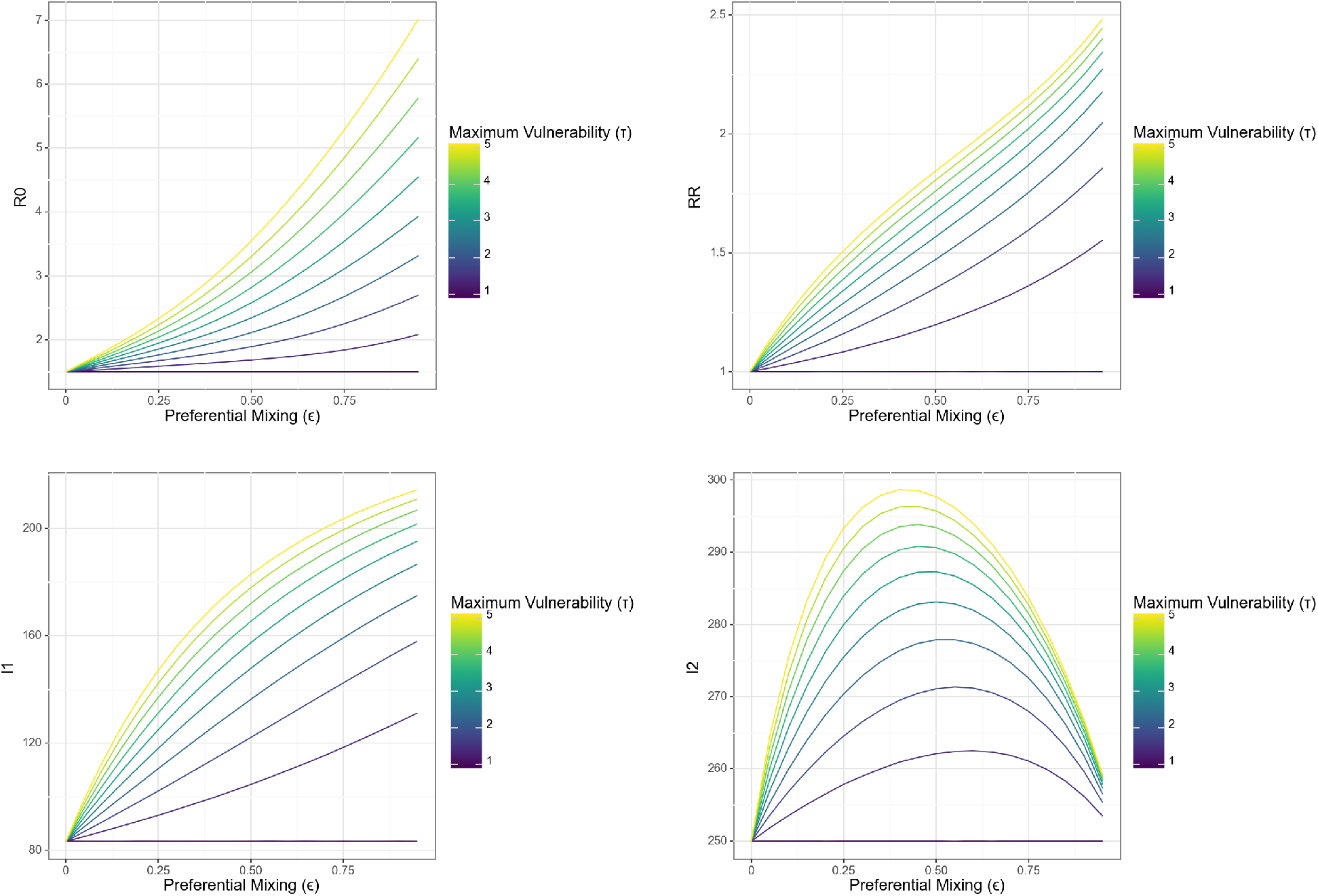
ℛ_0_ and relative risk of infection at equilibrium as a function of segregation intensity in fundamental cause model. The top left-hand panel illustrates the scaling of the population-level parameter ℛ_0_ as a function of increasing segregation. The top right-hand panel illustrates scaling of the relative risk of infection for the minority group vs. majority group as a function of increasing segregation. The bottom two panels represent the absolute prevalence in each group as a function of the model parameters. Note the curvilinear relationship between prevalence in the majority group with increasing segregation. Each colored line represents a different value of the maximum vulnerability, *τ* . For this figure, the parameters *β* = 1.5, *γ* = 1, *N*1 = 250 and *N*2 = 750 are fixed.

## 4. Discussion

In the analysis above, we have formalized the mental model relating residential segregation to infection inequities originally laid out by Acevedo-Garcia (24). The most important insight from interrogating this conceptual model is that spatial separation and vulnerability act interactively in the production of infection inequities associated at a high level with residential segregation. Specifically, as the degree of vulnerablity increases, the share of new infections among the minority group at any level of isolation grows, again illustrating how these mechanisms may work together to generate stark inequities in infection outcomes.

Figure 2 illustrates the role that preferential mixing and differential vulnerability may play in shifting risks away from the majority to the minority group: As population-wide risk increases with growing ℛ_0_, the share of new infections among the minority group grows rapidly. Notably, for small differences in *ρ*, this relationship is non-linear and rapidly grows with preferential mixing (*ϵ*).

When we examined vulnerability and separation as linked, correlated mechanisms driven by a high-level fundamental cause, structural racism, we see that it is easy to recreate the strong relationship between geographic separation and inequality that we might expect to see in the real world. However, our results show how examining this association alone can mask the role of differential vulnerability a key effect of segregation on infeciton inequities: As illustrated in Figure 2, even in the absence of spatial separation, differences in vulnerability to infection directly related to segregation, i.e. housing quality and crowding, healthcare access and comorbid illnesses, can generate large inequities in rates of infection. This is important, because the stereotypical understanding of residential segregation as a process resulting in extreme physical separation does not apply in all contexts.

For example, the alley houses inhabited by many African American residents of Jim Crow Baltimore (7) directly abutted the homes of comparatively better-off White residents. In addition, many alley house residents worked as servants and came into regular, direct, and sustained contact with the White residents of the street-facing homes. Conventional segregation measures, such as isolation and dissimilarity are likely to miss this type of relationship (35), and a simple regression of infection inequity on such a metric would likely show a null association. Similarly, a mathematical model in which the impacts of segregation are primarily operationalized via differential contact between and within groups would have little to say about these risks. By contrast, a multi-dimensional conceptualization of residential segregation as a ‘bundle of risks’, like the one we have presented here, can easily accomodate this understanding.

With that said, the model we examined in this analysis is - deliberately - a drastic simpliciation of a complex set of socio-biological problems. First, while the problem we discuss is highly spatial in nature, our model does not explicitly represent spatial relationships between groups or individuals. This was intentional, but necessarily limits the applicability of our model and findings to real-world data. Future elaborations on this modeling framework that include explicit spatial relationships will be important for understanding the generality of our key findings. In addition, our analysis focuses in on only one part of the picture of infection inequity. While segregation-associated drivers of exposure and infection are key drivers of infection inequity (13), differential risks of severe disease, death, and long-term sequelae of infection (e.g. long Covid) are also critically important manifestations of infection inequity which much be addressed by mathematical models.

Going forward, exploration of more-detailed theoretical and empirical models is critical for facilitating a deeper and principled integration of the theory and methods of infectious disease modeling with those of the social sciences. A particular area of need is for a more thorough theoretical and data-driven accounting of the nature of vulnerability in the context of infectious disease transmission. While in this analysis we have kept the concept of vulnerability broad and schematic, more detailed attention is necessary to make this concept useful for modeling, policy, and intervention.

This need is particularly urgent given the widespread use of summary metrics, such as the Social Vulnerability Index, to explain spatial, socioeconomic and racial differences in infection outcomes. While increased recognition of social vulnerability as a correlate of infection inequity is positive, when metrics such as SVI are treated as omnibus, black-box metrics, they can end up obscuring rather than drawing attention to the mechanisms that drive inequity in infection, illness and death (36). Figure 5 is a sketch of what a more comprehensive treatment of vulnerability in empirical and theoretical models of infection inequity might look like. Specifically, this conceptual model provides a view of how structural racism impacts specific dimensions of vulnerability and the downstream impacts of these sub-mechanisms on infection risk, severe disease, and death.

**Figure 5.**
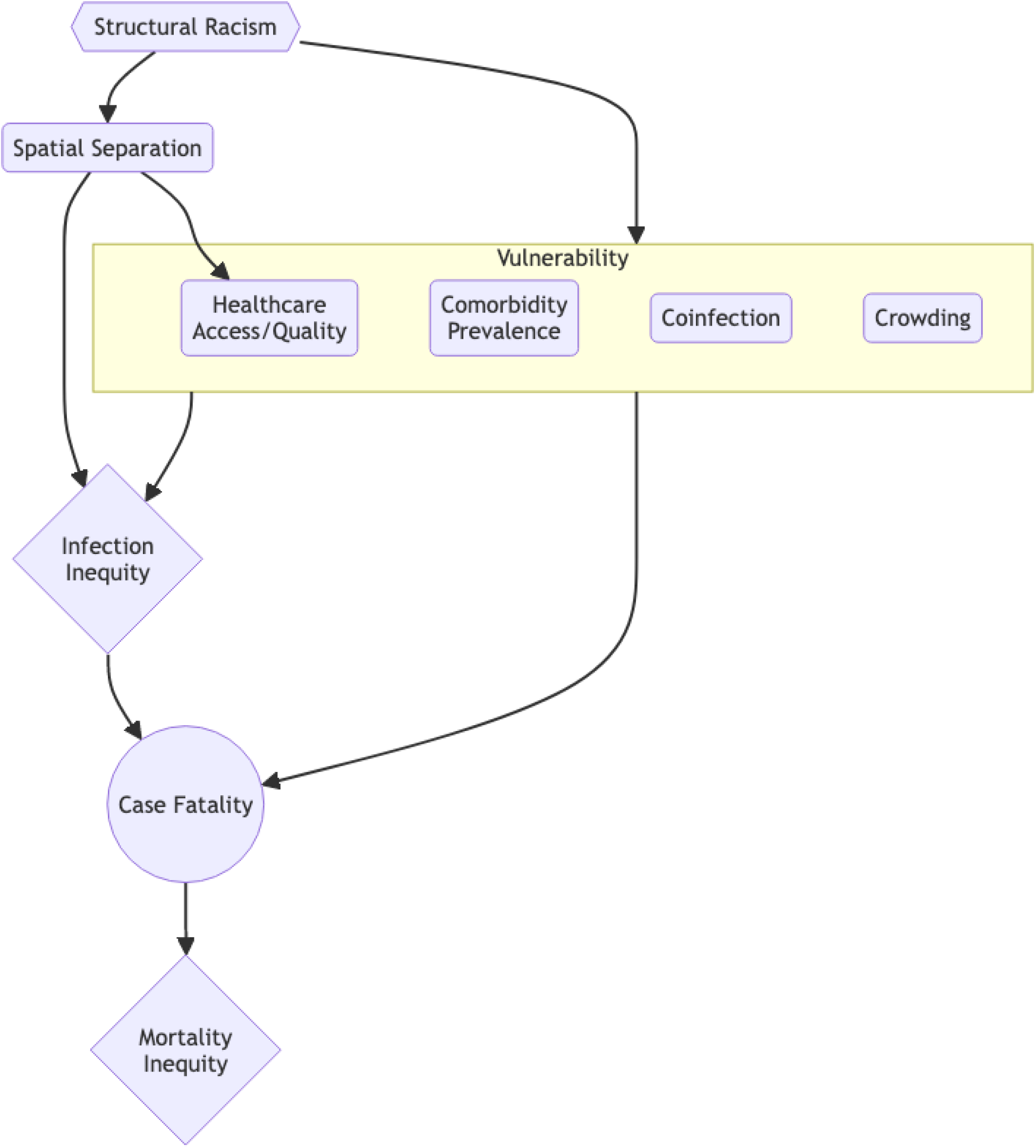
Expanded set of relationships between racism, residential segregation, vulnerability, and inequity in infection and death. This diagram illustrates specific factors that may be associated with vulnerability to infection and to poor infection outcomes, and their relationship to structural racism.

At the same time, the key insight of fundamental cause theory - that the effects of the high-level, structural causes is to put affected populations ‘at risk of risk’, and that this key function persists even when the intervening mechanisms change - underscores why it is important to resist the temptation to enumerate all possible intervening mechanisms linking structural racism and socioeconomic inequality to disparate infection outcomes. At the end of the day, it is the high-level fundamental causes that orchestrate most of the intermediary mechanisms, and an over-focus on these mechanisms can lead to ever-increasing model detail at the expense of the clear insights provided by FCT.

